# Subtyping Social Determinants of Health in *All of Us*: Network Analysis and Visualization Approach

**DOI:** 10.1101/2023.01.27.23285125

**Authors:** Suresh K. Bhavnani, Weibin Zhang, Daniel Bao, Mukaila Raji, Veronica Ajewole, Rodney Hunter, Yong-Fang Kuo, Susanne Schmidt, Monique R. Pappadis, Elise Smith, Alex Bokov, Timothy Reistetter, Shyam Visweswaran, Brian Downer

**Affiliations:** School of Public and Population Health, University of Texas Medical Branch, Galveston, TX, USA; Institute for Translational Sciences, University of Texas Medical Branch, Galveston, TX, USA; Division of Geriatric Medicine, Department of Internal Medicine, University of Texas Medical Branch, Galveston, TX, USA; College of Pharmacy and Health Sciences, Texas Southern University, TX, USA; Department of Population Health Sciences, Long School of Medicine, University of Texas Health San Antonio, San Antonio, TX, USA; School of Health Professions, University of Texas Health San Antonio, San Antonio, TX, USA; Department of Biomedical Informatics, University of Pittsburgh, Pittsburgh, PA, USA; Intelligent Systems Program, University of Pittsburgh, Pittsburgh, PA, USA

## Abstract

A.

**Background:** Social determinants of health (SDoH), such as financial resources and housing stability, account for between 30-55% of people’s health outcomes. While many studies have identified strong associations among specific SDoH and health outcomes, most people experience multiple SDoH that impact their daily lives. Analysis of this complexity requires the integration of personal, clinical, social, and environmental information from a large cohort of individuals that have been traditionally underrepresented in research, which is only recently being made available through the *All of Us* research program. However, little is known about the range and response of SDoH in *All of Us*, and how they co-occur to form subtypes, which are critical for designing targeted interventions.

**Objective:** To address two research questions: (1) What is the range and response to survey questions related to SDoH in the *All of Us* dataset? (2) How do SDoH co-occur to form subtypes, and what are their risk for adverse health outcomes?

**Methods:** For Question-1, an expert panel analyzed the range of SDoH questions across the surveys with respect to the 5 domains in *Healthy People 2030* (*HP-30)*, and analyzed their responses across the full *All of Us* data (n=372,397, V6). For Question-2, we used the following steps: (1) due to the missingness across the surveys, selected all participants with valid and complete SDoH data, and used inverse probability weighting to adjust their imbalance in demographics compared to the full data; (2) an expert panel grouped the SDoH questions into SDoH factors for enabling a more consistent granularity; (3) used bipartite modularity maximization to identify SDoH biclusters, their significance, and their replicability; (4) measured the association of each bicluster to three outcomes (depression, delayed medical care, emergency room visits in the last year) using multiple data types (surveys, electronic health records, and zip codes mapped to Medicaid expansion states); and (5) the expert panel inferred the subtype labels, potential mechanisms that precipitate adverse health outcomes, and interventions to prevent them.

**Results:** For Question-1, we identified 110 SDoH questions across 4 surveys, which covered all 5 domains in *HP-30*. However, the results also revealed a large degree of missingness in survey responses (1.76%-84.56%), with later surveys having significantly fewer responses compared to earlier ones, and significant differences in race, ethnicity, and age of participants of those that completed the surveys with SDoH questions, compared to those in the full *All of Us* dataset. Furthermore, as the SDoH questions varied in granularity, they were categorized by an expert panel into 18 SDoH factors. For Question-2, the subtype analysis (n=12,913, d=18) identified 4 biclusters with significant biclusteredness (Q=0.13, random-Q=0.11, z=7.5, *P*<0.001), and significant replication (Real-RI=0.88, Random-RI=0.62, *P<*.001). Furthermore, there were statistically significant associations between specific subtypes and the outcomes, and with Medicaid expansion, each with meaningful interpretations and potential targeted interventions. For example, the subtype *Socioeconomic Barriers* included the SDoH factors *not employed, food insecurity, housing insecurity, low income, low literacy*, and *low educational attainment*, and had a significantly higher odds ratio (OR=4.2, CI=3.5-5.1, *P*-corr<.001) for depression, when compared to the subtype *Sociocultural Barriers*. Individuals that match this subtype profile could be screened early for depression and referred to social services for addressing combinations of SDoH such as *housing insecurity* and *low income*. Finally, the identified subtypes spanned one or more *HP-30* domains revealing the difference between the current knowledge-based SDoH domains, and the data-driven subtypes.

**Conclusions:** The results revealed that the SDoH subtypes not only had statistically significant clustering and replicability, but also had significant associations with critical adverse health outcomes, which had translational implications for designing targeted SDoH interventions, decision-support systems to alert clinicians of potential risks, and for public policies. Furthermore, these SDoH subtypes spanned multiple SDoH domains defined by *HP-30* revealing the complexity of SDoH in the real-world, and aligning with influential SDoH conceptual models such as by Dahlgren-Whitehead. However, the high-degree of missingness warrants repeating the analysis as the data becomes more complete. Consequently we designed our machine learning code to be generalizable and scalable, and made it available on the *All of Us* workbench, which can be used to periodically rerun the analysis as the dataset grows for analyzing subtypes related to SDoH, and beyond.

## B. Introduction

Social determinants of health (SDoH), such as financial resources^1^ and housing stability,^2^ account for between 30-55% of people’s health outcomes.^3^ While many studies have identified strong associations among specific SDoH and health outcomes, most people experience multiple SDoH concurrently in their daily lives.^4–8^ For example, limited access to education, unstable employment, and lack of access to healthcare tend to frequently co-occur across individuals leading to long-term stress and depression.^8^ Such complex interactions among multiple SDoH make it critical to analyze combinations of SDoH versus single factors. However, analysis of such co-occurrences and their risks for adverse health outcomes requires the integration of personal, clinical, social, and environmental information, critical for designing cost-effective and targeted interventions. Unfortunately, the lack of databases containing such multiple datatypes from the same individuals has resulted in a fragmented understanding of how SDoH co-occur and impact health, critical for designing targeted interventions.

The *All of Us* program^9–11^ provides an unprecedented opportunity to address this fragmented view of SDoH. This program aims to collect data from multiple sources related to one million or more individuals with a focus on populations that have been traditionally underrepresented in biomedical research. These data sources include electronic health records (EHRs), health surveys, whole sequence genome data, physical measurements, and personal digital information. Critically, *All of Us* provides several survey modules containing a wide range of SDoH, which in combination with other data sources, could transform our understanding of high-risk combinations of SDoH.^9^

However, little is known about the range and response of SDoH in *All of Us*, and how they co-occur to form subtypes, which are critical for designing targeted medicine interventions. To address these gaps, we characterized 110 SDoH in *All of Us*, which guided the methods we used to analyze how they co-occur to form subtypes, and their risk for health outcomes. The results helped to highlight the opportunities and challenges for conducting subtype analysis in *All of Us*, which integrates multiple datatypes by using scalable and generalizable machine learning methods targeted to the design of targeted interventions.

## C. Background

### Social Determinants of Health

The World Health Organization (WHO) defines SDoH as the “non-medical factors that influence health outcomes.”^3^ Specifically, these include the conditions in which people are born, grow, work, live, and age. Furthermore, such conditions are shaped by a wider set of forces such as economic and social policies, and systems such as discriminatory laws and structural racism.

Several models have proposed the factors and mechanisms involved in SDoH.^4,12^ These models were motivated by the concept of *social gradient*,^13^ an empirical phenomenon observed within and across nations,^14,15^ consistently showing that the lower an individual’s social socioeconomic position, the worse their health. To help explain the factors underlying the social gradient, the Dahlgren-Whitehead model^4,16^ proposed several interconnected layers of social determinants that influence health. As shown in Fig. 1, the innermost layer contains demographic and genetic factors which are largely unmodifiable. In contrast, the outer layers are modifiable to different degrees such as lifestyle (e.g., exercise and smoking), social and community networks (e.g., contact with supportive friends and family), living and working conditions (e.g., access to health care and employment), and broader socio-economic, cultural, and environmental conditions (e.g., crime in the neighborhood). While this model was not intended to provide explicit testable hypotheses,^4^ the factors within each layer are expected to co-occur and impact each other, in addition to responding to external forces such as racism, and capitalism when it is focused on financial profits at the expense of societal benefits.

**Fig. 1.**
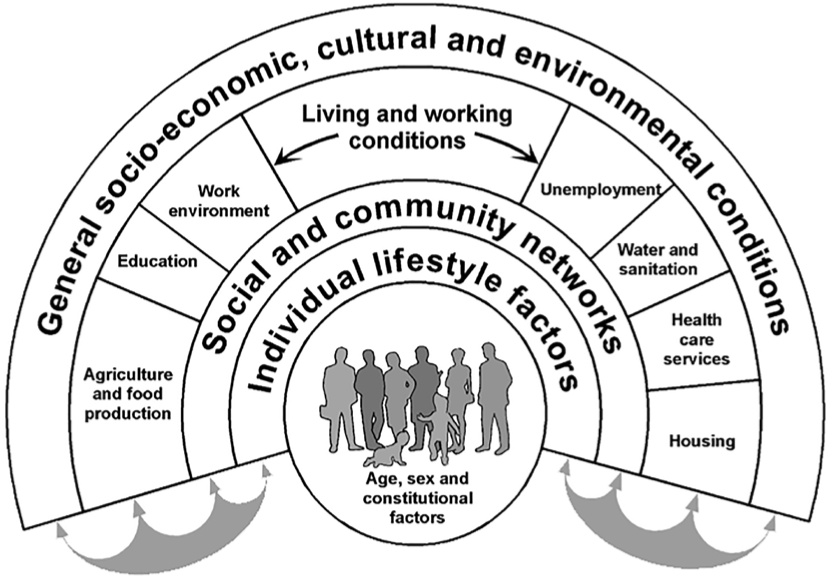
The Dahlgren-Whitehead conceptual model aimed to visually show the inter-related layers of SDoH domains that influence health.

These early SDoH models motivated numerous studies^17^ that analyzed associations among specific SDoH (e.g., immigration status and home density^7^), their association with health outcomes (e.g., education and mortality^18^), and how they manifest within subpopulations (e.g., patients with diabetes^19^). More recently, organizations such as Centers for Disease Control and Prevention (CDC) and *Healthy People 2030* (*HP-30*) have organized these empirical results into SDoH domains that roughly map to the Dahlgren-Whitehead model. For example, *HP-30* organizes SDoH empirical studies into five SDoH domains: (1) Economic Stability; (2) Education Access and Quality; (3) Health Care Access and Quality; (4) Neighborhood and Built Environment; and (5) Social and Community Context. Furthermore, the PhenX program (that provides well-established measurement protocols for use in biomedical and translational research) has identified SDoH data collection protocols to enable more systematic data collection and analysis.^20–22^

While the above findings and categorizations have greatly improved our understanding of SDoH and their impact on health, they have been mostly analyzed based on snapshots of associations among a few factors and health outcomes. In contrast, SDoH models and recent empirical studies suggest that multiple SDoH tend to co-occur and impact each other. For example, during the pandemic, Hispanic and Black or African American individuals not only had a higher exposure to COVID-19 infections due to their front-line jobs and overcrowded living conditions, but also had a higher risk for serious infections due to prior conditions not addressed due to lack of healthcare access.^4^ Similarly, undocumented immigrants with lower incomes living in neighborhoods with high pollution, combined with the stress of deportation, have an increased risk of multiple chronic conditions such as depression and lung cancer.^7^ Such studies have resulted in the Centers for Medicare and Medicaid Services (CMS) emphasizing that SDoH are a multi-level construct which includes both individual and contextual factors that have complex interactions.^23^

The above co-occurrences of multiple SDoH and their impact on health directly reflect the interconnected layers of the Dahlgren-Whitehead shown in Fig. 1. However, analysis of such co-occurrences and their health outcomes requires large datasets with multiple datatypes that have only recently been made available through the *All of Us* program.

### *All of Us*: Multiple Datatypes Across a Large Cohort of Underrepresented Americans

The *All of Us* research program^9–11^ (*All of Us*), funded by the National Institutes for Health since 2015, aims to accelerate biomedical research to enable discoveries leading to individualized and equitable prevention and treatment. Such research is currently hampered due to the *limited range* of personal, clinical, social, and environmental variables available for the same individuals, *limited representation* in research datasets of socially marginalized populations, and *limited access* to individual-level data due to privacy laws.

To overcome these hurdles, *All of Us* provides three critical features: (1) a data repository that is projected to contain one million or more participants, with data from multiple sources including electronic health records (EHRs), health surveys, whole sequence genomic data, physical measurements, and personal digital information such as from Fitbits; (2) a cohort targeted to include 75% participants from populations underrepresented in research (race, ethnicity, gender, sex, sexual orientation, and disability) oversampled from the US population; and (3) strictly-enforced rules to prevent reidentification of participants by disallowing the download of any participant data, or reporting research results for subgroups less than 20. These rules allow analysis of the *All of Us* data to be categorized as non-human subjects research, which combined with training and personal authentication by researchers, has resulted in a substantial reduction in administrative hurdles.

As of 12/30/22 (Controlled Tier, version 6), *All of Us* contained 372,397 total participants, with 8.6% who had attempted all 9 health surveys (7 related to demographics and general health, and 2 related to COVID-19), and 26.5% who had genomic data. Critical to the current study is the recent addition of a survey specifically targeted to SDoH questions, which has been attempted by 15.5% in the *All of Us* cohort. A preliminary analysis revealed that SDoH appear to be distributed across multiple health surveys and EHR codes, with participants providing those data at different times on a rolling basis. However, little is known about the range and response of SDoH in *All of Us*, and how they co-occur to form subtypes, a critical step for selecting the methods to identify and interpret SDoH subtypes.

### Computational Methods to Identify and Interpret Subtypes

A wide range of studies^24–32^ on topics ranging from molecular to environmental determinants of health have shown that most humans tend to share a subset of characteristics (e.g., comorbidities, symptoms, genetic variants), forming distinct subtypes (also referred to as *subgroups* or *subphenotypes* depending on the condition and variables analyzed). A primary goal of precision medicine is to identify such subtypes and infer their underlying disease processes to design interventions targeted to those processes.^25,33^ Methods to identify subtypes include: (a) investigator-selected variables such as race for developing hierarchical regression models,^34^ or assigning patients to different arms of a clinical trial, (b) existing classification systems such as the Medicare Severity-Diagnosis Related Group (MS-DRG)^35^ to assign patients into a disease category for purposes of billing, and (c) computational methods such as classification^36–38^ and clustering^28,39^ to discover subtypes.

Several studies have used computational methods to identify subtypes, each with critical trade-offs. Some studies have used *combinatorial* approaches^40^ (identify all pairs, all triples etc.), which are intuitive, but which can lead to a combinatorial explosion (e.g., enumerating combinations of the 31 Elixhauser comorbidities would lead to 2^31^ or 2147483648 combinations), with most combinations that do not incorporate the full range of symptoms (e.g., the most frequent pair of symptoms ignores what other symptoms exist in the profile of patients with that pair). Other studies have used *unipartite* clustering methods^38,39^ (clustering patients or comorbidities, but not both together) such as k-means, and hierarchical clustering; and dimensionality-reduction methods such as principal component analysis (PCA) to help identify clusters of frequently co-occurring comorbidities.^40–46^ However, such methods have well-known limitations including the requirement of inputting user-selected parameters (e.g., similarity measures, and the number of expected clusters), in addition to the lack of a quantitative measure to describe the quality of the clustering (critical for measuring the statistical significance of the clustering). Furthermore, because these methods are unipartite, there is no agreed-upon method to identify the patient subgroup defined by a cluster of variables, and vice-versa.

More recently, bipartite network analysis^47^ (see Appendix A for additional details) has been used to address the above limitations by automatically identifying *biclusters,* consisting of patients and characteristics simultaneously. This method takes as input any dataset such as *All of Us* participants and their SDoH, and outputs a quantitative and visual description of biclusters (containing both participant subgroups, and their frequently co-occurring SDoH). The quantitative output generates the number, size, and statistical significance of the biclusters,^48–50^ and the visual output displays the quantitative information of the biclusters through a network visualization.^51–53^ Bipartite network analysis therefore enables (1) the automatic identification of biclusters and their significance, and (2) the visualization of the biclusters critical for their clinical interpretability. Furthermore, the attributes of participants in a subgroup can be used to measure the subgroup risk for an adverse outcome, to develop classifiers for classifying a new participant into one or more of the subgroups, and to develop a predictive model that uses that subgroup membership for measuring the risk of an adverse outcome for the classified participant.

However, while several studies^50,54–61^ have demonstrated the usefulness of bipartite networks for the identification and clinical interpretation of subgroups, there has been no systematic attempt to identify SDoH subtypes mainly because of the lack of large cohorts containing a wide coverage of SDoH. The *All of Us* program provides an opportunity to use bipartite networks for the identification and interpretation of SDoH subtypes using a wide range of variables in a large cohort, and for analyzing their risk for health outcomes, a critical step in advancing precision medicine.

## D. Method

### Research Questions

Our analysis was guided by two research questions targeting the *All of Us* dataset:

1. *What is the range and response to survey questions related to SDoH?*
2. *How do SDoH co-occur to form subtypes, and what are their risk for adverse health outcomes?*

### Expert Panel

The selection of the research questions, variables, cohort, methods, results, and their interpretation were guided by an expert panel consisting of SDoH researchers with a professional background in applied demography, gerontology, and rehabilitation, who worked closely with the machine learning and biostatistics researchers. The overall project and manuscript were examined by an ethicist for bias, stigma, and perpetuation of stereotypes. The examination of each step in the project is therefore aligned with the human-centered artificial intelligence approach.^62–64^

### Data Description

#### Study Population

In Question-1, we analyzed the full *All of Us* cohort (n=372,397) and characterized their responses to all the SDoH identified by the expert panel (described in the Variables subsection). For Question-2, we analyzed all participants (n=12,913) that had valid responses to the SDoH identified in Question-1, and used them to identify subtypes, and their risks for specific outcomes.

#### Variables

For Question-1, the expert panel was asked to review all 1113 questions across 7 *All of Us* non-COVID health surveys, each of which is attempted once per participant (*The Basics, Lifestyle, The Basics, Personal Medical History, Health Care Access & Utilization, Family Health History, and SDoH*), and the 2843 Systematized Nomenclature of Medicine (SNOMED) codes related to SDoH.^65^ The expert panel arrived at a consensus for the SDoH across the surveys and the SNOMED codes. As the SDoH-related SNOMED codes in the EHR had very low usage (see Appendix B for a characterization), they were not further characterized.

In Question-2, to identify and analyze the SDoH subtypes, we used the following variables:

- Independent variables included the SDoH factors identified from Question-1.
- Covariates including 3-digit zip code (to determine if participants in each subtype came from a state that accepted Medicaid expansion providing greater access to health insurance), and demographics (age, sex, race).
- Outcomes included: (1) *<u>Depression</u>* was selected as it is a common health outcome when individuals encounter SDoH in their daily lives such as long-term stress resulting from racism,^66^ and dysregulation of the hypothalamic-pituitary-adrenal axis (HPA) axis.^67^ Depression was defined as having a positive response to both of the following questions in the *The Basics* survey (“*Are you still seeing a doctor or health care provider for depression?*” and “*Has a doctor or health care provider ever told you that you have Depression?*”) or having SNOMED codes related to depression Codes in their EHR (35489007, 36923009, 370143000, 191616006, or 66344007), (2) *<u>Delayed Medical Care</u>* was selected as it often results from the lack of medical insurance, which can impact the use of medical care when needed leading to poorer health outcomes.^68^ Delayed medical care was defined as having one or more positive responses to 9 survey questions (delayed care due to: transportation, rural, nervousness, work, childcare, copay, elderly care, out of pocket, and deductible) from the *Health Care Access* & *Utilization* survey. (3) *<u>Emergency Room (ER) Visits in Last Year</u>* was selected because lack of medical insurance often results in individuals not seeking early medical care when needed, leading to an exacerbation of conditions precipitating one or more ER visits.^69^ As the survey questions that we used for SDoH subtyping were based on outcomes in the past year, we defined ER visits for a participant as having one or more ER visits (CPT 99281-99285) one year preceding the date when the SDoH survey was completed.

### Analytical Approach

***Question-1:*** What is the range and response to survey questions related to SDoH?

To address this question, we characterized all SDoH in *All of Us* at two levels of granularity: (1) SDoH questions based on the surveys used to collect the data, and (2) SDoH factors, which were categories of the SDoH questions to form a coarser grained classification (see Table-1 which explains SDoH questions, factors, and subtypes). These two levels of SDoH granularity in *All of Us* were characterized as follows:

**Table 1.**
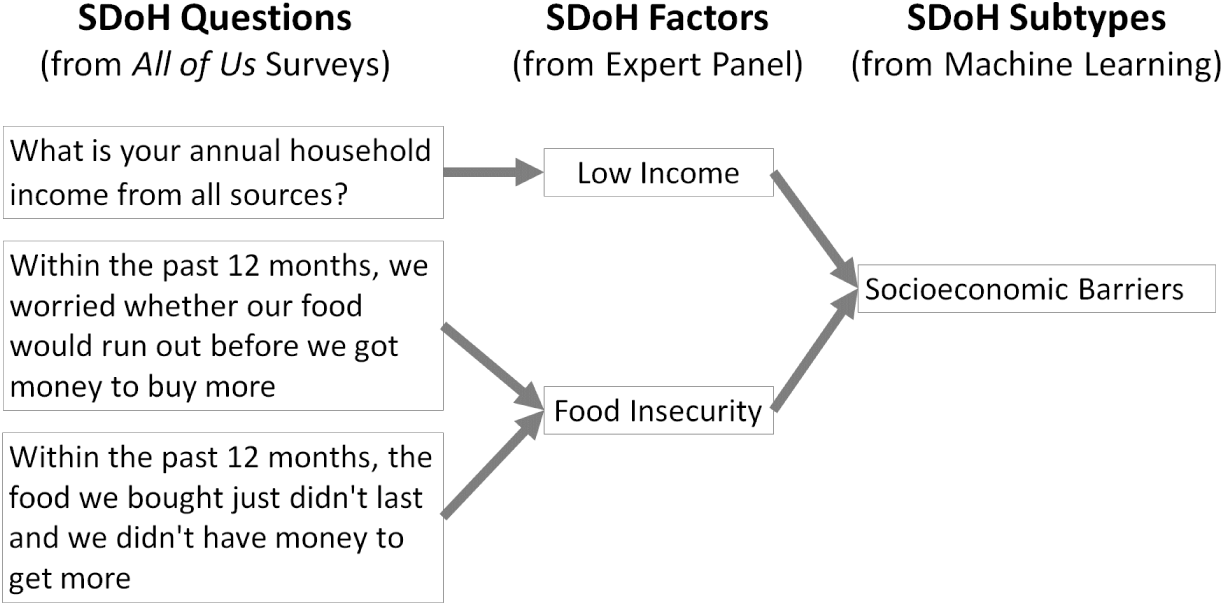
Examples showing how the <u>SDoH questions</u> from the *All of Us* surveys which differed in their levels of granularity, were transformed by the expert panel into <u>SDoH factors</u> with uniform granularity to ensure consistency for analysis and interpretation, and clustered into <u>SDoH subtypes</u> through machine learning. The SDoH questions and factors were subsequently analyzed for coverage across the 5 HP-30 domains (see Appendix C for more details).

#### Identification and Coding of SDoH (SDoH Questions and SDoH Factors)

##### A. Identification and Coding of SDoH Questions in All of Us

Members of the expert panel independently used their domain knowledge about SDoH to identify and code the SDoH questions, and to examine their range with respect to the five *HP-30* domains using the following steps: (1) reviewed all 1113 questions across 7 health surveys (excluding 2 related to COVID-19), and extracted all SDoH questions that were relevant; (2) transformed all positive or value-free questions into negative phrases and abbreviated them for interpretability in the graphs (e.g., “*How often do you have someone help you read health-related materials?*” was changed into *“No one to help read health materials”*); (3) reverse coded, and dichotomized the abbreviated SDoH questions (e.g., Always/Often=1, and Never/Occasionally/Sometimes=0); and (4) categorized the SDoH questions into one of the five *HP-30* SDoH domains (Economic Stability, Education Access and Quality, Health Care Access and Quality, Neighbourhood and Built Environment, and Social and Community Context). The expert panel subsequently met and collaboratively resolved any differences between their coding schemes to arrive at a consensus (see Appendix-C for the 110 SDoH questions, and their consensus coding by the expert panel).

##### B. Identification and Coding of SDoH Factors

The expert panel arrived at a consensus to categorize one or more of the above SDoH questions in *All of Us,* into SDoH factors, and to examine their range with respect to *HP-30* using the following steps: (1) reviewed the subgrouping labels of questions in the *All of Us* surveys, and integrated them to categorize the SDoH into factors; (2) coded a participant as having a “1” for a SDoH factor if they had one or more of the questions within that factor which had been answered with a “1”; and (3) categorized the SDoH factors into one of the five *HP-30* SDoH domains (Economic Stability, Education Access and Quality, Health Care Access and Quality, Neighbourhood and Built Environment, and Social and Community Context) (see Appendix-C for the 110 SDoH questions, their consensus coding into 19 SDoH factors, and mapping to the 5 SDoH domains from HP-30).

### Range and Responses to SDoH Questions and Factors

The above knowledge-based classification of SDoH questions and SDoH factors were analyzed to examine their range (with respect to the five *HP-30* domains), and their response (across all participants in *All of Us*), using the following methods. (1) Bar graph displaying the number of participants that had valid answers (all responses other than “skip” or “choose not to answer”) to each of the SDoH questions, sorted by survey based on mean response, and then sorted by raw response within each survey. Additionally, to examine their range, each SDoH question/factor was colored by one of the five SDoH domains defined by *HP-30*. (2) Venn diagram showing how many participants had cross-sectionally valid responses to all identified SDoH questions/factors. (3) Table describing the number and proportion of race, ethnicity, sex, gender, and age between those that answered the SDoH questions/factors, versus those that did not have valid responses. (4) Frequency distribution of the number of SDoH questions/factors across participants that had valid responses for all the SDoH questions. The above plots are shown in the Results section.

***Question-2:*** How do SDoH co-occur to form subtypes, and what are their associations with covariates and risks for adverse health outcomes?

#### Data

We used the cohort identified in Question-1 (participants who had valid answers to all the SDoH questions). However, *e*xamination of the SDoH questions revealed that some of them (e.g., cannot afford dental care, cannot afford prescriptions) had a finer level of granularity compared to others (e.g., single household). As the questions with a finer level of granularity tend to be more strongly co-related to each other in comparison to other coarser grained questions, they also tend to cluster together more strongly, confounding the interpretation of the subtypes. In contrast, as the SDoH factors had a more uniform granularity, and were at a level of abstraction that was appropriate to guide referral to the proper social services, we used them to identify the SDoH subtypes.

#### Analytical Model

To identify SDoH subtypes, their associations with outcomes and covariates, and their future translation into precision medicine, we used a three-part analytical framework called **H**eterogenization, **I**ntegration, and **T**ranslation (HIT). As shown in Fig. 2, the *heterogenization* step was used to identify the subtypes through the use of bipartite modularity maximization^48–50^ (see Appendix A for more details), the *integration* step was used to measure the association of each subtype to multiple datatypes,^70^ and the *translation* step was used to qualitatively interpret the subtypes,^70^ with the goal of developing in the future a decision-support system to translate the subtypes into clinical practice. The following describes the specific methods used in each of the HIT steps:

**Fig. 2.**
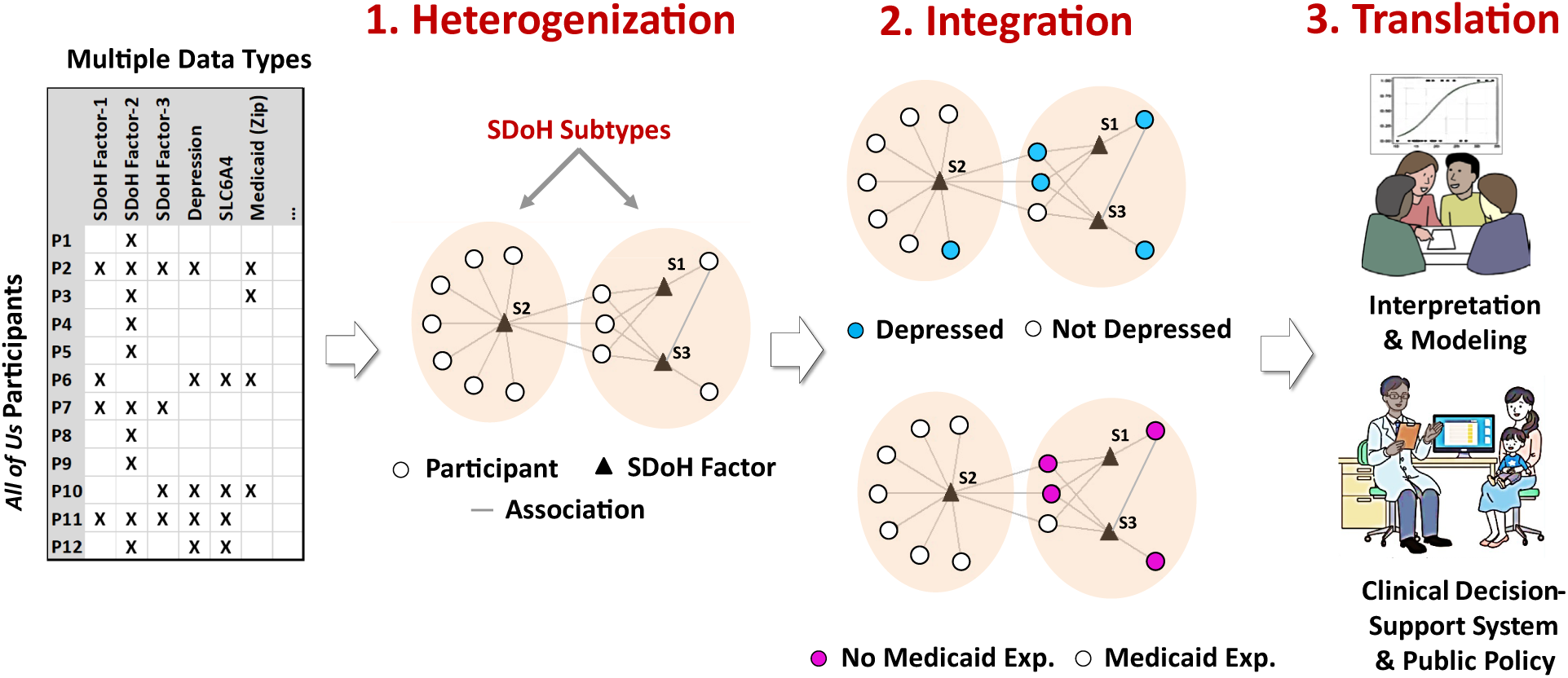
The three steps of the **HIT** framework to analyze SDoH. (1) **Heterogenization** of the data to identify subtypes. (2) **Integration** of multiple datatypes such as from EHRs (e.g., depression), and state (e.g., to determine Medicaid expansion) to determine risk and enrichment of each subtype, and (3) **Translation** of subtypes through interpretation and predictive modeling, with the goal of designing clinical decision-support systems and public policy.

##### 1. Heterogenization: Identification of Subtypes

As there were many participants that did not have valid answers to the SDoH questions, dropping them resulted in differences in the proportion of demographic variables compared with the full *All of Us* cohort. The data therefore needed to be adjusted to better reflect the overall *All of Us* participants. To adjust the demographic distribution of the cohort to match the full *All of Us* cohort, we calculated the inverse probability weights (IPW)^71,72^ for each participant in our cohort. IPW calculates weights to proportionally boost the values of participants that are underrepresented in our cohort, with respect to a comparison such as the full *All of Us* data, using the method similar to an earlier study of *All of Us*^73^ (see Appendix E). Next, we multiplied the IPW generated weights with the original binary values for each participant in our cohort, and used *min-max* to range-normalize those weights within each SDoH factor. Finally, to test the replicability of the SDoH factor biclustering, we randomly divided the dataset into a training and a replication dataset.

We identified subtypes in the training dataset, and tested the degree to which the SDoH factor co-occurrences replicated in the test dataset using the following steps: (1) modelled participants and SDoH factors as a weighted bipartite network (see Step-1 in Fig. 2) where nodes were either participants (circles), or SDoH factors (triangles), and the associations between participant-SDoH factor pairs were weighted edges (lines) generated from IPW. The inclusion of IPW generated weights enabled the network to represent the demographic distribution of the full *All of Us* data; (2) used a bipartite modularity maximization algorithm,^48–50^ (which takes edge weights into consideration) to identify the number of biclusters, their members, and measure the degree of biclusteredness through bicluster modularity (Q, defined as the fraction of edges falling within a cluster, minus the expected fraction of such edges in a network of the same size with randomly assigned edges); (3) measured the significance of Q by comparing it to a distribution of the same quantity generated from 1000 random permutations of the network, while preserving the network size (number of nodes), and the distribution of weighted edges for each participant; (4) used the Rand Index (RI) to measure the degree to which SDoH occurred and did not co-occur in the same cluster in the training and test datasets,; and (5) measured the significance of RI by comparing it to the mean of a distribution of the same quantity generated by randomly permuting the training and replication datasets 1000 times, while preserving the size of the networks.

##### 2. Integration: Risk and Enrichment of Subtypes

We used logistic regression to measure the odds ratio (OR) for each subtype compared pairwise to each of the other subtypes, for the three outcomes (Depression, Delayed Medical Care, and ER Visits in Last Year), and for living in a state with Medicaid expansion. To adjust for the difference in demographics due to the missingness, we used weights generated from IPW for each participant, and the comparisons were adjusted for demographics (age, sex, race) and corrected for multiple testing within each outcome using FDR. As 1688 (13.1%) participants did not have 3-digit zip code information, we used IPW to measure the weights of the cohort, and used them to account for potential sample selection bias.

##### 3. Translation: Interpretation of Subtypes

The subtype interpretation was done using the following steps: (a) used the *Fruchterman-Reingold* ^51^ and *ExplodeLayout*^52,53^ algorithms to visualize the bipartite network along with the risk for each of the outcomes; (b) asked the expert panel to independently label the subtypes, infer the mechanisms that increase the risks in each subtype for the three outcomes (Depression, Delayed Medical Care, and ER Visits in Last Year) with potential strategies to reduce those risks, and then collaboratively come to a consensus; and (c) asked an ethicist to examine the results and their interpretations for bias, stigma, and perpetuation of stereotypes.

## E. Results

***Question-1:*** What is the range and response to survey questions related to SDoH?

### Identification and Coding of SDoH Questions and Factors

The expert panel identified 110 questions from 4 surveys (*The Basics*, *Overall Health*, *Healthcare Access & Utilization, and SDoH)*. Of these, 110 were abbreviated, and 48 were negatively-worded and coded (see Appendix C). The 110 SDoH questions were further categorized into 19 SDoH factors (one of these was *Delayed Medical Care* that was used as an outcome).

### Response to SDoH Questions and Factors

As shown in Figure 3A, the number of valid responses for each of the 110 SDoH questions was largely dictated by the surveys in which they were solicited. SDoH from 2 surveys (*The Basics*, *Overall Health*) had the most valid responses (mean=349434, SD=23556), followed by *Healthcare Access & Utilization* (mean=149898, SD=6146), and finally the *SDoH* survey (mean=55960, SD=1083). This pattern of responses matched how answers to each of the surveys were solicited: at enrollment, all participants are required to do *The Basics*, and *Overall Health* surveys, and then on a rolling basis the other surveys responses are solicited. The *SDoH* survey is the latest survey that was solicited, which explained their lowest number of responses. As shown in Fig. 3B, this pattern of missingness held for the responses at the SDoH factor level, which was not unexpected as the SDoH factors were aggregations of the SDoH questions. However, as shown in Fig. 3A and 3B by the uneven number of valid responses within each survey block, there were several SDoH questions that had invalid responses (“skip” or “chose not to answer”) at both levels of granularity: *The Basics*: 6%; *Health Access & Utilization* 6.1%; *Overall Health*: 4.39%; and *SDoH*: 2.61%. Furthermore, the proportion of valid to invalid responses between them was significantly different for the SDoH questions (χ^2^ (2, N=365237)=57.489, *P*<.001), and for the SDoH factors (χ^2^ (2, N=372063)=75.637, *P<.001*).

**Fig. 3.**
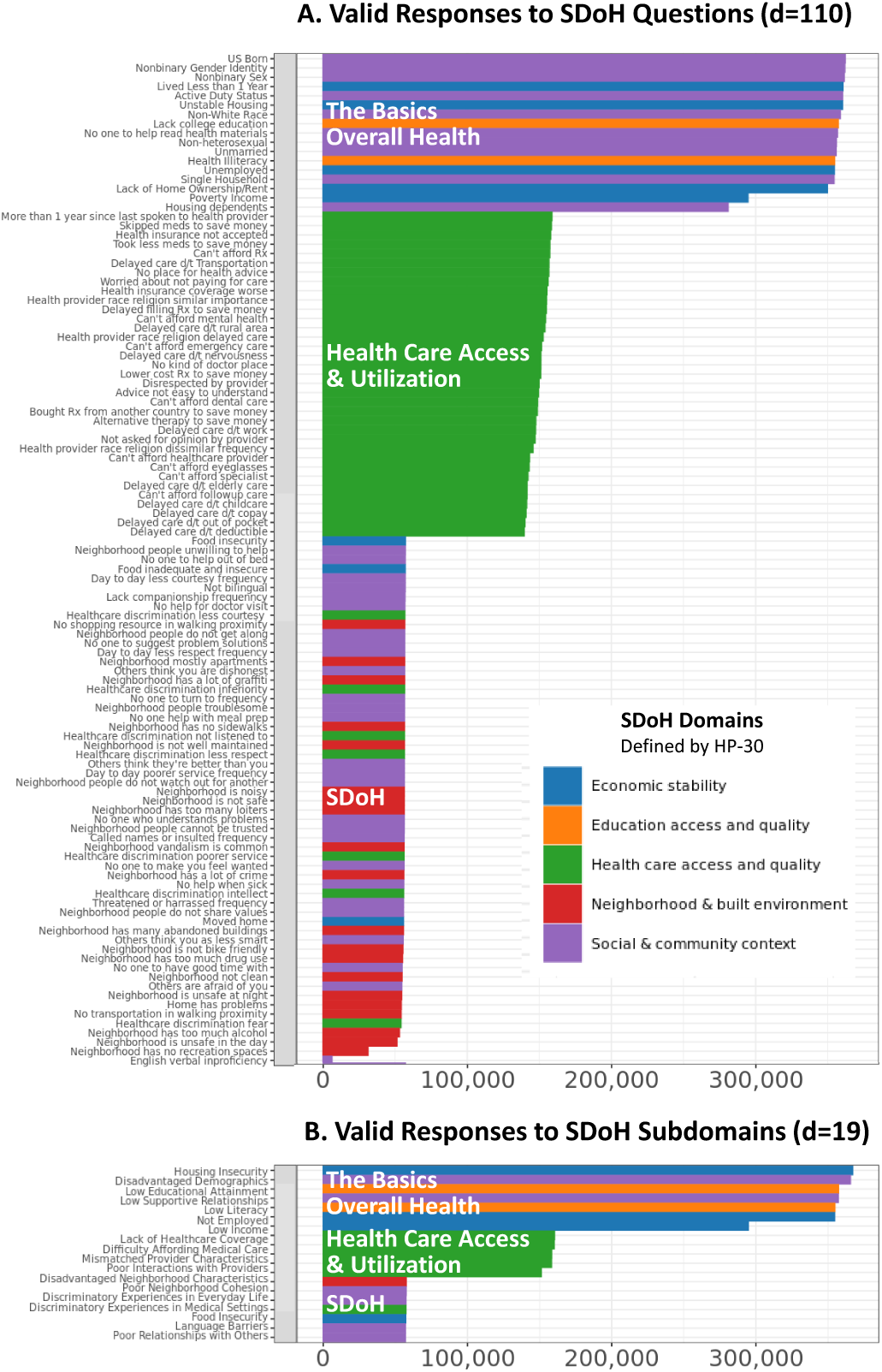
The number of valid responses for (A) 110 SDoH questions, and (B) 19 SDoH factors. The colors denote how the SDoH in each were categorized based on the 5 *HP-30* domains.

### Range of SDoH Questions and Factors

As shown by the colored bars in Figure 3, the surveys spanned the full range of the five SDoH *HP-30* domains. The SDoH questions in *The Basics* and *Overall Health* surveys were predominantly related to economic stability (blue) and social and community context (purple), those in *Healthcare Access & Utilization* survey were all related to that topic (green), whereas those from the *SDoH* survey were a mix of all four domains. Overall, the four surveys contained 110 SDoH questions that together had 100% coverage of the five *HP-30* domains: Social and Community Context=38; Neighborhood and Built Environment=19; Economic Stability=10; Education Access and Quality=2; Health care Access and Quality=42. This characterization suggests that while the SDoH in *All of Us* have broad domain coverage across the surveys, analysis of them requires access to all four surveys, each of which have different levels of completion and valid responses.

### Cohort with Maximized Valid Responses

Given the large degree of missingness in 2 of the 4 surveys, we could not use multiple imputation to estimate the values. We therefore had to find a subset of participants that had valid responses to all the SDoH questions. An examination revealed that two SDoH questions had <10% responses (*English Verbal Frequency*=1.67%, and *Neighborhood has no recreation spaces*=8.4%), accounting for the largest loss in cohort size with valid responses. These questions were therefore dropped from further analysis. Furthermore, one question required a branched response (*Living Situation* branching to *Did not Live in a House*) which were merged. Finally, as we used *Delayed Medical Care* as an outcome, 9 questions related to that topic were removed, resulting in a total of 98 SDoH questions. As shown in Fig. 4, a Venn diagram of the overlap among the valid responses across the surveys revealed that 12,913 participants had valid responses to all 98 SDoH questions.

**Fig. 4.**
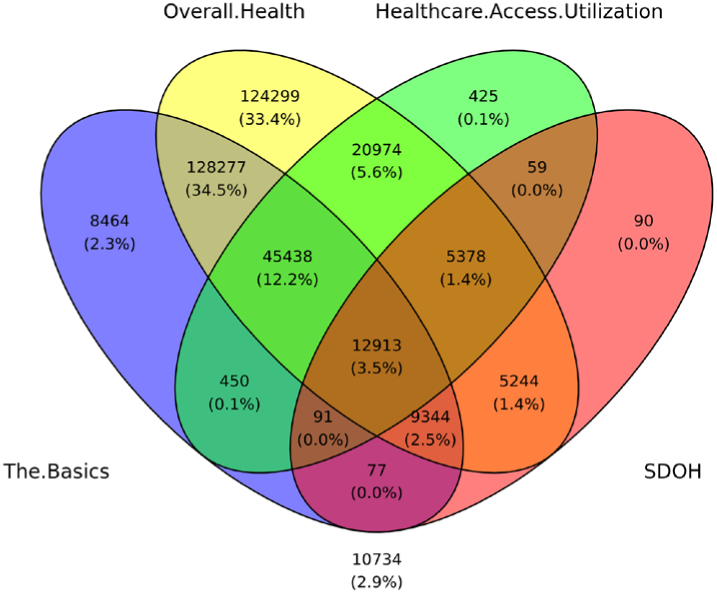
Venn diagram showing 12,913 participants (3.5% of the full cohort), who had valid responses to all 98 SDoH questions.

### Co-occurrence of the Number of SDoH across Responders

As shown in Fig. 5, participants had a median of 15 SDoH question co-occurrences and a median of 9 SDoH factors co-occurrences. Furthermore, participants

**Fig. 5.**
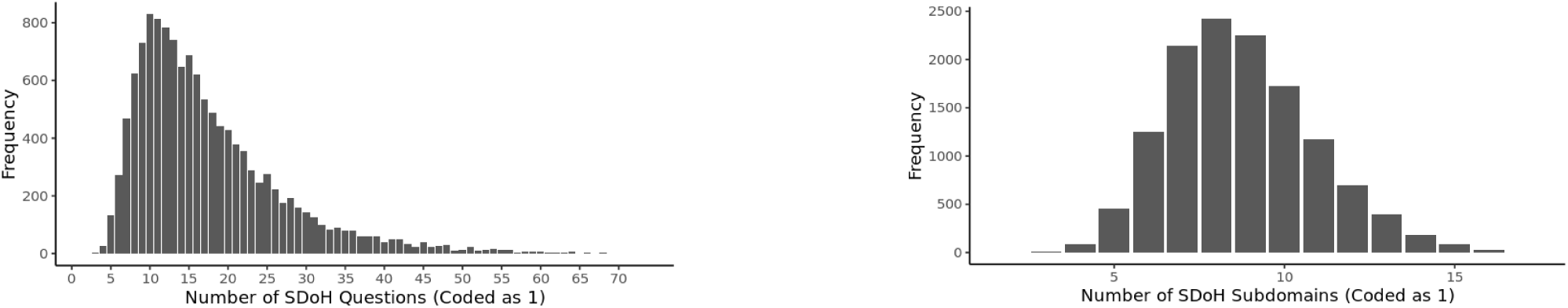
Frequency distribution of (a) number of co-occurring responses to **SDoH questions** across the 12,913 participants with valid answers to the 98 SDoH questions, and (b) number of co-occurring **SDoH factors** across 19 SDoH factors.

of color or racial/ethnic minorities, who had valid responses to the 110 SDoH questions, had a significantly higher median number of co-occurring SDoH compared to the equivalent White population (median participants of color or racial/ethnic minorities=20, median White=14, *P*<.001). These results show the high co-occurrences of SDoH at both levels of granularity, with a significant difference in median co-occurrences between the White and the participants of color or racial/ethnic minority populations, with valid responses.

### Participant Demographics with Valid Responses to SDoH Questions

As the cohort size dropped to 3.5%, we analyzed how that impacted the demographic distribution compared with the overall *All of Us* data. As shown in Table 2, there were statistically significant differences in race (χ^2^(5, *N*=372,397)=2073.1, *P*<.001), and ethnicity (χ^2^(9, *N*=372,397)=6292.2, *P*<.001) between the two cohorts, after multiple testing correction, with a higher proportion of White participants having valid answers compared to participants of color, or racial or ethnic minorities. Furthermore, there was a statistically significant difference in age between the participants who had valid answers, versus those that did not (*H*(1)=148.08, *P*<.001). These results show the demographic differences between the cohort with complete and valid answers to the SDoH questions, in comparison to the full *All of Us* data, necessitating the need for weights generated from IPW to address those imbalances.

**Table 2.** The demographic differences between the total *All of Us* participants, and those that had valid answers to all 110 SDoH questions. ^a^Participants that completed all questions, and did not skip, or choose not to answer a question; ^b^Age 122 = a participant chose the least birth year (1900). Participant counts less than 20 are shown as a count of 20 based on the *All of Us* reporting rules.

| Demographics |  | All AoU Participants:<br>372,397 (100%) | All AoU Participants with<br>valid<sup>a</sup> SDoH answers:<br>12,913 (3.5%) |
| --- | --- | --- | --- |
| Race | White | 201149 (54.01%) | 11279 (87.35%) |
|  | Black or African American | 73383 (19.71%) | 482 (3.73%) |
|  | Asian | 12459 (3.35%) | 324 (2.51%) |
|  | Other or >1 population | 26890 (7.22%) | 343 (2.66%) |
|  | None Indicated | 58516 (15.71%) | 485 (3.76%) |
| Ethnicity | Not Hispanic or Latino | 288227 (77.4%) | 12095 (93.67%) |
|  | Hispanic or Latino | 66704 (17.91%) | 751 (5.82%) |
|  | Additional Options | 17466 (4.69%) | 67 (0.52%) |
| Sex at birth | Female | 222495 (59.75%) | 8236 (63.6%) |
|  | Male | 138831 (37.28%) | 4674 (36.09%) |
|  | Intersex | 80 (0.02%) | 20 (0.15%) |
|  | Additional Options | 10991 (2.95%) | 20 (0.15%) |
| Gender | Female | 220833 (59.3%) | 8113 (62.82%) |
|  | Male | 138140 (37.09%) | 4642 (35.95%) |
|  | Non Binary | 920 (0.25%) | 60 (0.46%) |
|  | Transgender | 464 (0.12%) | 20 (0.15%) |
|  | Additional Options | 12040 (3.23%) | 79 (0.61%) |
| Age |  | Median=56 (19-122<sup>b</sup>) | Median=58(19-93) |

***Question-2:*** How do SDoH factors co-occur to form subtypes, and what are their risk for adverse health outcomes?

The cohort used to identify the subtypes consisted of 12,913 participants, of which 12,886 had valid IPW weights. The latter cohort were split randomly into the training and replication datasets, each with complete data for 18 SDoH factors (identified in Question-1), in addition to the three outcomes (depression, delayed medical care and ER visits in last year), and covariates (demographics).

### 1. Heterogenization: Identification of Subtypes

The subtypes were identified by using a bipartite network where the edges were weighted using the IPW generated weights to account for the imbalance in demographics between our cohort and the full *All of Us* data. The weighted bipartite network of the training dataset (n=6492) and the 18 SDoH factors revealed 4 biclusters with statistically significant bicluster modularity (Q=0.13, random-Q=0.11, z=7.5, *P*<0.001). As shown in Fig. 6, there were four clusters with participant subgroups and their most frequently co-occurring SDoH factors (*<u>Cluster-1</u> (pink)*: low education attainment, low literacy, low income, not employed, food insecurity, and housing insecurity; *<u>Cluster-2</u> (green)*: difficulty affording medical care, discriminatory experiences in everyday life, discriminatory experiences in medical settings, poor interactions with providers; *<u>Cluster-3</u> (blue)*: poor neighborhood cohesion, and poor relationships with others; and *<u>Cluster-4</u> (gray)*: disadvantaged demographics, language barriers, lack of healthcare coverage, mismatched provider characteristics, disadvantaged neighborhood characteristics, and low supportive relationships). These co-occurrences of SDoH factors, significantly replicated in the replication data set (Real-RI=0.88, Random-RI=0.62, *P<*.001). As shown in Fig. 7, while the 18 SDoH factors have a hierarchical relationship with the five *knowledge-driven HP-30* domains (shown on the left), those same SDoH factors have a more complex relationship with the four *data-driven* biclusters (shown on the right).

**Fig. 6.**
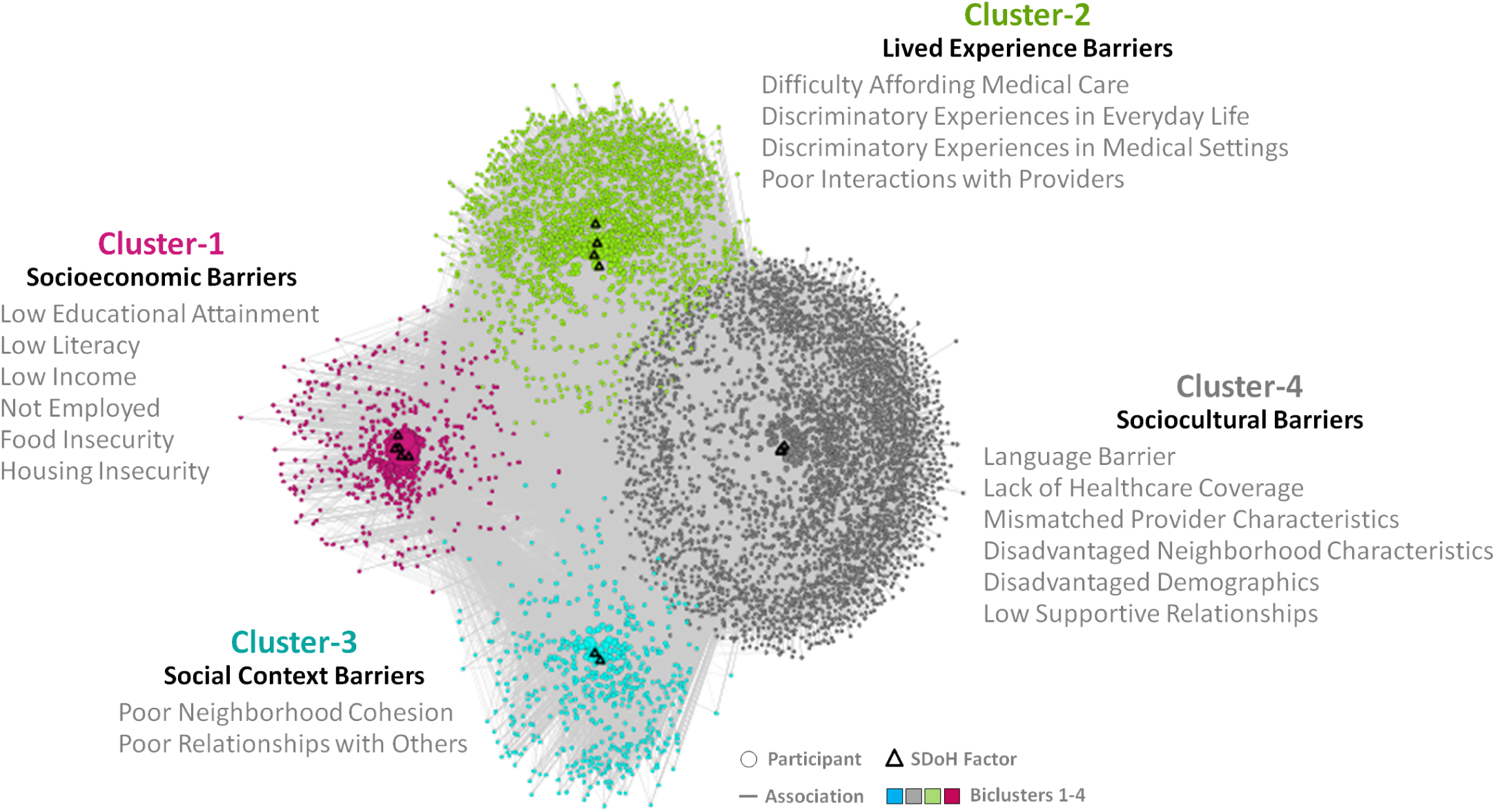
Four biclusters in the training dataset consisting of subgroups of participants (n=6492), and their most frequently co-occurring SDoH factors (d=18) (see Appendix B for SDoH questions related to the SDoH factors clustered within each subtype). The biclustering was significant (Q=0.13, random-Q=0.11, z=7.5, *P*<0.001) and the co-occurrence of the SDoH factors significantly replicated in the replication dataset (Real-RI=0.88, Random-RI=0.62, *P<*.001). Across all three outcomes, Cluster-1 had a significantly higher OR compared to Cluster-4. The cluster labels in bold text represent the consensus interpretion by the expert panel.

**Fig. 7.**
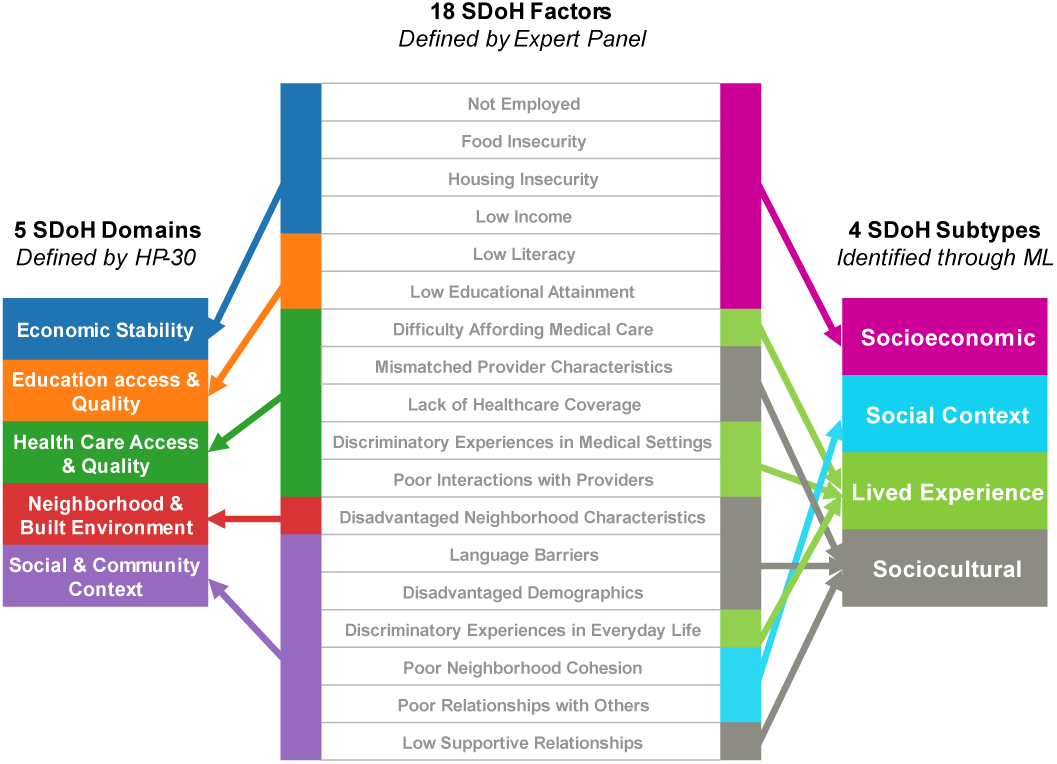
18 SDoH factors (center) have a hierarchical relationship with the 5 SDoH domains define by HP-30 (left), both of which are knowledge driven. In contrast, the SDoH factors have a complex relationship with the SDoH subtypes (right) identified through machine learning (ML), reflecting how they co-occur in the real-world, and aligned with models such as the Dahlgren-Whitehead model (shown in Fig. 1).

### 2. Integration: Risk and Enrichment of Subtypes

Table 3 shows the association of each subtype to the three outcomes. As shown by the dark orange row, Cluster-1 (low educational attainment, low literacy, low income, not employed, food insecurity, and housing insecurity) had a significantly higher OR for each of the three outcomes compared to Cluster-4 (mismatched provider characteristics, disadvantaged neighborhood characteristics, lack of healthcare coverage, disadvantaged demographics, low supportive relationships, language barrier). Furthermore, within the *Depression* outcome, each of the clusters had a significantly higher OR compared to one other cluster forming a ranking of risk among all the four clusters (1>3>2>4). In contrast, *Delayed Medical Care* had two other significant associations (2>1, 3>4), with *ER Visit in the Last Year* having only the one significant pair-wise association that fit into the overall trend.

**Table 3.** Across all three outcomes, Cluster-1 had a significantly higher risk compared to Cluster-4 (dark orange row). The Depression outcome had a distinct ranking of risks (light orange), whereas the other two outcomes had a subset of them.

| Cluster Comparison |  | Outcomes |  |  |
| --- | --- | --- | --- | --- |
| Cluster-A vs. Cluster-B |  | Depression | Delayed Medical Care | ER Visit in Last Year |
| 1 | 2 | OR=1.7, CI=1.5-2, P-corr=2.5e-10 <.001 | OR=0.78, CI=0.67-0.92, P-corr=0.0038 <.01 | OR=1.2, CI=0.91-1.6, P-corr=0.24 |
| 1 | 3 | OR=1.3, CI=1.1-1.6, P-corr=0.019 <.05 | OR=0.88, CI=0.72-1.1, P-corr=0.23 | OR=1.4, CI=0.96-1.9, P-corr=0.13 |
| 1 | 4 | OR=4.2, CI=3.5-5.1, P-corr=3.5e-52 <.001 | OR=3.5, CI=3-4.1, P-corr=1.8e-53 <.001 | OR=1.8, CI=1.4-2.3, P-corr=0.00016 <.001 |
| 2 | 3 | OR=0.79, CI=0.64-0.97, P-corr=0.022 <.05 | OR=1.2, CI=0.98-1.4, P-corr=0.094 | OR=1, CI=0.75-1.5, P-corr=0.8 |
| 2 | 4 | OR=2.3, CI=1.9-2.7, P-corr=5.2e-21 <.001 | OR=4.3, CI=3.7-5, P-corr=3.4e-85 <.001 | OR=1.3, CI=1-1.7, P-corr=0.12 |
| 3 | 4 | OR=2.9, CI=2.3-3.5, P-corr=1.5e-21 <.001 | OR=3.6, CI=3-4.4, P-corr=1.6e-38 <.001 | OR=1.4, CI=0.95-1.9, P-corr=0.13 |

As shown in Table 4, this trend continued in the enrichment analysis of association with living in a state with *No Medicaid Expansion*. As shown, Cluster-1 had a significantly higher OR compared to Cluster-4, in addition to the other clusters. The overall results suggest that Cluster-1 and Cluster-4 form “book ends” representing the high and low ends of risk among the clusters.

**Table 4.** Cluster-1 had a significantly higher OR compared to Cluster-4 (dark orange) for no Medicaid expansion, in addition to Cluster-2 and Cluster-3 (light orange).

| Cluster Comparison |  | Enrichment |
| --- | --- | --- |
| Cluster-A vs. Cluster-B |  | No Medicaid Expansion |
| 1 | 2 | OR=1.5, CI=1.3-1.8, P-corr=1.7e-05 <.001 |
| 1 | 3 | OR=1.3, CI=1-1.6, P-corr=0.048 <.05 |
| 1 | 4 | OR=1.3, CI=1.1-1.5, P-corr=0.0057 <.01 |
| 2 | 3 | OR=0.99, CI=0.8-1.2, P-corr=0.97 |
| 2 | 4 | OR=0.99, CI=0.86-1.2, P-corr=0.97 |
| 3 | 4 | OR=1, CI=0.82-1.2, P-corr=0.97 |

### 3. Translation: Interpretation of SDoH Subtypes and Design of Potential Interventions

The expert panel examined the co-occurrences of SDoH factors within each bicluster shown in the network visualization (Fig. 6), and integrated them with the quantitative ORs in Table 3 and 4. The consistent “book ends” result where Cluster-1 had significantly higher ORs compared with Cluster-4 across all four variables was of strong interest, and interpreted as follows: (1) **Cluster-1** was labeled *Socioeconomic Barriers* as it contained multiple high risk SDoH. These co-occurring SDoH could have resulted from cascades over time such as low educational attainment, potentially leading to lower rates of employment and lower income, with higher rates of food and housing insecurity. Such cascading factors can be perceived as being relatively unmodifiable, leading to a higher risk for chronic stress and depression. Furthermore, the strong association of this subtype with the outcomes *Delayed Medical Care* and *ER Visits in Past Year*, and that participants in this subtype were more likely to be from a US state with *No Medicaid Expansion*, provided a more comprehensive understanding of this high-risk SDoH subtype. (2) **Cluster-4** was labeled *Sociocultural Barriers* as it contained a combination of SDoH related to disadvantaged neighborhood characteristics, and low supportive relations, in addition to language barriers, and mismatched provider interactions. In contrast to socioeconomic barriers in Cluster-1, many of the sociocultural barriers could be perceived as potentially modifiable, resulting in a lower risk for depression, delayed medical care, and ER visits. Participants that match this profile could be screened for language and communication barriers, useful for providing culturally-competent care, identifying providers that better match the profile of the individuals, and for providing resources to facilitate contact with matching nationality or cultural groups online or in the vicinity.

While Cluster-1 and Cluster-4 formed the “book ends” of risk across the three outcomes potentially caused by relative differences in the unmodifiability of their frequently co-occurring SDoH, **Cluster-2** was flagged as critical and labeled *Lived Experience Barriers*. The SDoH in this cluster included discriminatory experiences in everyday life and in medical settings, in addition to poor interactions with providers and difficulty in affording medical care. These frequently co-occurring SDoH could explain why this subtype had a significantly higher OR for *Delayed Medical Care* compared to Cluster-1. Finally, **Cluster-3** was labeled *Social Context Barriers* as the SDoH related to poor neighborhood cohesion and relationships with others. While not as critical as Cluster-1 and Cluster-2, this cluster still had significantly higher OR for depression compared to Cluster-4. Together, the four clusters could explain how different degrees of unmodifiability in frequently co-occurring SDoH might impact health outcomes.

The expert panel and the ethicist concluded that clinicians treating patients that match each subtype profile could be alerted of specific risks, and consequently motivate a discussion about mental health and consequences of delayed medical care, with the goal of collaboratively exploring options and solutions with the patients. The results could also be useful for resource planning in hospitals to ensure there was adequate staff to address the needs of populations they serve, and for proposing public policies to address the critical connection between specific combinations of SDoH, and their impact on public health.

Furthermore, the subtypes did not have a one-to-one mapping to the 5 SDoH domains defined by *HP-30*. As shown in Fig. 7, these data-driven clusters have a complex relationship with the SDoH domains and factors. While one subtype belonged to a single domain (subtype *Social Context* belonged to the domain *Social and Community Context*), three of the four subtypess belonged to two or more domains (e.g., the subtype *Socioeconomic Barriers* belonged to the domains *Economic Stability*, and *Education Access and Quality*). Such interdomain relationships reflect how SDoH co-occur in the real world reflecting the complex cross-domain interactions described in the Dahlgren-Whitehead model (Fig. 1). These relationships could be useful for refining conceptual models to explain the complex association beween SDoH and adverse health outcomes, and to build more accurate SDoH models for predicting adverse health outcomes.

## F. Discussion

The mechanisms through which SDoH precipitate adverse health outcomes are complex consisting of many interacting factors and feedback loops among individual and environmental/contextual factors. While this phenomenon has been studied for more than three decades, critical hurdles for researchers have included the *limited range* of data types, *limited representation* of populations that have been socially marginalized, and *limited access* to individual-level data at scale due to privacy laws. Recognizing that *All of Us* has well-articulated plans and resources to overcome these limitations, but is still in a rapidly evolving stage, we conducted a systematic characterization of more than a hundred SDoH available in *All of Us,* and used them to identify SDoH subtypes with the future goal of designing targeted interventions. This attempt led to the following opportunities and challenges related to data, methods, and theory.

### Data: Missingness and Granularity

*All of Us* data contained 110 SDoH across 4 surveys, and 93 SDoH-related SNOMED codes in the EMRs. While these provided a comprehensive coverage of SDoH with respect to domains and factors identified by *HP-30*, our analysis uncovered the following patterns of missingness and SDoH granularity.

#### Missingness

The analysis revealed three types of missingness: (1) *Rollout Missingness:* This type of missingness was largely dictated by how the surveys were rolled out to participants. As all participants at enrollment are required to do *The Basics*, and *Overall Health* surveys, they had the highest responses, followed by the later solicited surveys *Healthcare Access & Utilization*, and *SDoH* rolled out more recently in 2022. This order of rollout was the main source of missingness resulting in a precipitous reduction in cohort size for those that had answers to all the SDoH questions. (2) *Valid Answer Missingness.* As participants can choose not to answer any survey questions, the data contained “PMIs” related to “skip” and “choose not to answer”. These accounted for a much smaller reduction in cohort size for complete data. (3) *Low Usage Missingness.* Although there were 259 SDoH SNOMED codes, only 93 (3.3%) had such information for >20 participants that are allowed to be reported. This could be because most clinicians currently do not screen for SDoH, as it is typically done by the social worker. Furthermore, we also attempted to use 3-digit zip codes to determine which subtypes had a significant association to living in a state that did not offer Medicaid expansion. However, 13.1% (1688) of the participants did not have zip code information (which was adjusted by using IPW).

Together, the above three types of missingness impacted the size of the resulting cohort that had valid answers, in the following two ways: (1) a drastic reduction in cohort size by 93.5%. However, because of the size of the overall data (n=372,397), we were still left with a large cohort (n=12,886), which to the best of our knowledge is the largest set of individuals to be analyzed for such a wide range of SDoH; and (2) significant differences in the proportion of race, ethnicity, and age in the above cohort when compared to the overall *All of Us* population. Specifically, the cohort with valid answers had significantly more White, or non-Hispanic, or older participants, when compared to the overall cohort. This could potentially be because once a participant has been enrolled, there is a 90-day delay in sending subsequent solicitations to complete surveys, a policy that is currently being re-assessed due to its impact on missingness. We therefore had to correct this imbalance in demographic proportions by using IPW, with the goal of identifying subtypes that were representative of the overall *All of Us* cohort.

#### Granularity

Because our goal was to use machine learning methods to identify SDoH subtypes, we encountered uneven granularity in the SDoH questions. Some questions were fine-grained and highly correlated and therefore would cluster more strongly because of the nature of the granularity of the questions, not because of the SDoH mechanisms. To address this uneven granularity, and to make the results more interpretable, we used SDoH factors which had a coarser but more consistent level of granularity. We chose this approach because SDoH factors had already been defined, were understood by the expert panel enabling high domain fidelity, and appeared to be at the right level of abstraction useful for clinical applications such as referring a patient to the appropriate social services. However, because the use of coarse-grained variables loses information, future research could explore aggregating only those SDoH questions that are highly correlated, while preserving the rest at the finer level of granularity, and explore computational methods to merge SDoH questions into SDoH factors.

### Method: Scalability, Generalizability, and Extensibility

We designed the HIT analytical framework to be scalable enabling its use for the growing size of the data in *All of Us*, to be generalizable across cohorts and conditions, and to be extensible for including additional methods as needed in the future. Testing the HIT framework on the *All of Us* data provided insights for the strengths and limitations of the framework, and for the *All of Us* workbench where the analysis was conducted.

#### Scalability

We used three types of code to conduct the analysis for both research questions. (1) Automatically generated code to extract the cohort, produced by *All of Us* once a cohort was selected using the point and click interface. This code was adequately scalable and generalizable and so will not be discussed further. (2) Customized code to extract specific parts of the data. For example, the analysis of co-occurrences required customized code in R to plot the diagrams in Fig. 3. As expected, these tasks required strong programming skills, but fortunately we did not encounter any coding or execution problems using the R or Python programming languages. However, there were significant server issues which hampered our analysis. Although the workbench instructions state that code running on the workbench for more than 2 weeks would be terminated and all intermediate results deleted, we frequently encountered our work disappearing at shorter intervals. These disruptions resulted in a higher consumption of the free server time credits, and fewer analyses that we could conduct due to the computation time. (3) Machine learning code we had previously developed and disseminated on CRAN^74–76^ to conduct the bipartite network analysis and the significance testing, and to visualize the network. As this code was designed to be generalizable and scalable, we did not encounter any issues in the execution of our code (besides the same server issues mentioned above). Finally, the visualization of our networks worked as expected, and we used them to help interpret the patterns in the data.

#### Generalizability

Our code for the first two steps of the HIT framework is in Jupyter notebooks and have been used to analyze other cohorts that were filtered for age and prior conditions. For example, we extracted a cohort (n=4090) of participants with diabetes aged >=65 with complete data on 18 SDoH variables selected through consensus by 2 experienced health services researchers, and guided by Andersen’s behavioral model. The analysis^77,78^ revealed 7 SDoH subtypes with statistically significant modularity compared with 100 random permutations of the data (*All of Us*=.51, Random Mean=.38, z=20, *P*<.001), and which were not only clinically meaningful, but also significant in different degrees for the outcome. Our subsequent attempt at increasing the number of SDoH variables from 18 to 110 for participants with diabetes that had valid answers, led to an extremely small cohort size (n=926) (see Appendix D) due to the missingness that we described above. While this reduction resulted in our current strategy of analyzing all particpants regardless of condition or age, these experiments demonstrate that our approach is generalizable to other subsets of the data.

#### Extensibility

The HIT model is designed to be extensible to include other methods. For example, the model could use other biclustering (e.g., Non-negative Matrix Factorization^79^) and causal modeling methods, and use different types of classification (e.g., deep learning^80^), and prediction methods (e.g., subgroup-specific modeling ^38^) to build the decision-support system in the Translational Step (Fig. 2). Furthermore, the model can integrate a wide range of data types to enable analysis of how each subtype is associated with them, resulting in a layered interpretation of the SDoH subtypes as we have demonstrated. For example, as the percentage of participants that have genomic information increases (currently more than 25% of our cohort had missing genomic information), our pipeline will be able to integrate such information into our analysis. Finally, the integration of different datatypes required a diverse team consisting of experts in machine learning, biostatistics, programming, clinical care, health services research, gerontology, and ethics to enable a 360 analysis and interpretation of the subtypes, and therefore aligned with the human-centered artificial intelligence approach.^62–64^ Furthermore, the use of the workbench to share results through visualizations of the results operationalized *team-centered informatics*^81^ designed to facilitate multidisciplinary translational teams^82^ to work more effectively across disciplinary boundaries, with the goal of analyzing subtypes, and designing targeted interventions.

### Theory: Model Building, and Translational Implications

The identification of SDoH subtypes has strong implications for model building in addition to translational applications. As shown in Fig. 7, while the current classification of five SDoH domains has a hierarchical relationship with the SDoH factors, the data-driven clusters have a more complex association with the same SDoH factors. This reflects the complexity of how SDoH occur in the real-world, while at the same time being interpretable for purposes of translation.

Future models should develop predictive models using the data-driven subtypes to determine whether they improve the accuracy of predicting adverse health outcomes when compared to models that do not use those subtypes. Because the subtypes were clinically interpretable, they could be used to build classification and predictive models, and used with an interface to develop a clinical decision-support system that help to triage patients to critical services. For example, the St. Vincent House (https://www.stvhope.org/) in Galveston, Texas provides several services to address SDoH including free walk-in clinical care, nurse practitioner with small co-pay requested, English and Spanish-speaking free mental health counseling, free dental health clinic, utility and rental assistance, case management, financial literacy, expanded food pantry, weekly free home delivery of pantry groceries, snack pack for people experiencing homeless, free transportation for doctor’s appointment, immigration legal services, and spiritual counseling. Given the availability of this wide range of services in many communities across the US, a decision-support system could help to classify an individual based on their SDoH profile into one or more of the subtypes, measure their risk for an adverse health outcome. Such information could be used by clinicians to collaboratively explore solutions with the patient to consider more of such local services based on the membership strength for a subtype, and the associated risk (Fig. 2, Step-3). At a population level, understanding health risks associated with clusters may assist institutions and organizations in developing more effective prevention programs.

### Notebooks for *All of Us* Community Use

Because the missingness in SDoH variables is expected to reduce, their characterization and subtyping will need to be repeated and verified for different cohorts. Therefore, we have made the following two sets of code available for general use by *All of Us* researcher community (accessible after creating a free account on *All of Us* and completing the required training):

1. *SDoH Valid Answer Tracker.* This set of notebooks generate four plots which can be used by other researchers on *All of Us* to characterize any cohort: (1) valid responses plot to show how many participants have data with valid responses, and colored by SDoH domains; (2) Venn diagram showing how many participants have valid responses for all questions within each survey; (3) frequency distribution plot showing co-occurrence of SDoH across the selected cohort. This set of tools should enable researchers to characterize SDoH across different cohorts, to help determine methods that are appropriate to adjust for missingness in those cohorts.
2. *SDoH Subtyper.* This set of notebooks can be used to conduct the following analyses: (1) bicluster modularity of a cohort with the 18 SDoH factors to identify the number and members of biclusters, and the measure Q representing the quality of the biclustering; (2) visualization of the bipartite network; and (3) significance of the network with respect to null models.

### Limitations

This study has two main limitations. The first emerges from the temporary limitations of the large amount of missingness in the survey data, precluding the use of imputation methods which assume a random distribution of missingness. We could therefore use only complete data, which led to a large drop in cohort size, and which also introduced a bias in the demographics requiring a rebalancing through IPW. While such rebalancing is typically done for large datasets, the IPW method requires judgement to decide which variables to include in the model, and therefore could have introduced additional unknown biases. Therefore, the model should be refined to determine which variables to include in the regression models that estimate the IPWs. However, because the clustering was similar between the unweighted and IPW weighted networks, we believe that the current subtypes are stable, meaningful, and represent the demographic composition of the full *All of Us* data, but which needs to be verified by redoing the analysis as the data becomes more complete. The limitation of missingness in the surveys is expected to be addressed as *All of Us* has recently removed the requirement of waiting for 90 days before a subsequent survey is given to an enrollee in the program, potentially reducing the degree of missingness. The second limitation is due to the high computational cost of empirically determining the significance of the biclustering. As such analysis is computationally expensive and time-consuming, it limited the experiments we could do to test different cohorts and models. We therefore look forward to the *All of Us* workbench providing the ability to run batch processes more efficiently, and which will be uninterrupted for extended periods of time (exceeding the current time window), which together could help alleviate this computational hurdle in the future.

## G. Conclusion

How SDoH impact health is a complex phenomenon involving many interconnected social, biological, and environmental factors which have yet to be fully elucidated. While this phenomenon has been studied for more than 30 years, the analyses have been hampered by the lack of large cohorts representing diverse populations with a wide range of SDoH variables measured, multiple datatypes, and with easy access by researchers. *All of Us* provides an unprecedented opportunity to directly address these limitations with the goal of doing justice to early conceptual models such as the social gradient and the Dahlgren-Whitehead model, both of which drew international attention to the complex ways in which individual and contextual SDoH factors impact health. The *All of Us* dataset is also timely because of the extensive health disparities that were revealed during the pandemic, which highlighted the critical need to address SDoH in the public and policy realms. However, because *All of Us* is still rapidly evolving to meet its target of one million participants or more, we conducted a systematic characterization of SDoH variables in *All of Us,* and used the results to guide the analysis of SDoH subtypes. The subtypes identified along with their risks could be used to design data-informed interventions, resource planning strategies, and public health policies aimed towards reducing the risks for adverse outcomes. Careful consideration would be required to ensure that the identification of high-risk subtypes is not used in a way that stigmatizes subpopulations.

Our first goal of characterizing the data revealed the nature of the missingness in SDoH, and the uneven granularity in the SDoH questions. Both these results led us to select the IPW method to address the missingness, and analysis of subtypes at the SDoH factor level of granularity. Our second goal of identifying SDoH subtypes led not only to statistically significant biclusters, but also to their statistically significant replication, and meaningful domain interpretations. These results set the stage for further investigations to build and evaluate classification and prediction models for designing decision-support systems that alert clinicians of specific risks their patients face due to a combination of SDoH factors. The results also led to the design, use, and dissemination of general-purpose tools currently available on *All of Us* for other researchers, which will be useful to reanalyze the *All of Us* data as it grows over the next few years to directly address the high rate of missingness. These collaborative advances should position *All of Us* to revolutionize research for analyzing complex phenomena such as how SDoH impact health and beyond, with the goal of enabling a more equitable future that all of us deserve.

## Data Availability

All data analyzed in this study is available to researchers after training from the All of Us research program.

https://www.researchallofus.org/data-tools/workbench/

## H. Acknowledgments

The authors thank Gautam Vallabha for his assistance in refining the analysis and the manuscript. This study was supported in part by the Clinical and Translational Science Award (UL1 TR001439) from the National Center for Advancing Translational Sciences at the National Institutes of Health, the University of Texas Medical Branch Claude D Pepper Older Americans Independence Center funded by the National Institute of Aging (NIA) at the National Institutes of Health (P30AG024832), MD Anderson Cancer Center, and the National Library of Medicine (R01 LM012095) at the National Institutes of Health, and by the NIA (K01AG058789). The content is solely the responsibility of the authors and does not necessarily represent the official views of the National Institutes of Health.

## Appendix A

Description of Bipartite Network AnalysisA network consists of nodes and edges; nodes represent one or more types of entities (e.g., participants or SDoH), and edges between the nodes represent a specific relationship between the entities. Figure 1A shows a unipartite network where nodes are the same type (typically used to analyze co-occurrence of comorbidities^46^). In contrast, Figure 1B shows a bipartite network where nodes are of two types, and edges exist only between different types such as between participants (circles) and SDoH (triangles). Bipartite network analysis takes as input any dataset such as *All of Us* participants and their SDoH, and automatically outputs a quantitative and visual description of biclusters (containing both participant subgroups, and their frequently co-occurring SDoH). The quantitative output provides the number, size, and statistical significance of the biclusters,^48–50^ and the visual output displays the quantitative information of the biclusters through a network visualization.^51–53^ Bipartite network analysis therefore enables (1) the automatic identification of biclusters and their significance, and (2) the visualization of the biclusters critical for their clinical interpretability including labeling the subtypes, inferring potential mechanisms that precipitate adverse outcomes in each subtype, and designing targeted interventions to prevent them. Furthermore, the characteristics (e.g., outcomes and covariates) of participants in a subtype can be used to measure the risk of a subtype for an adverse outcome when compared to a reference group (e.g., a control group or another subtype), and therefore enables the integration of multiple data types. Finally, the biclusters can be used to develop classifiers for classifying a new participant into one or more of the subtypes, and developing a predictive model that uses those subtype membership for measuring the risk of an adverse outcome for that new participant.^70^

**Fig. 1.**
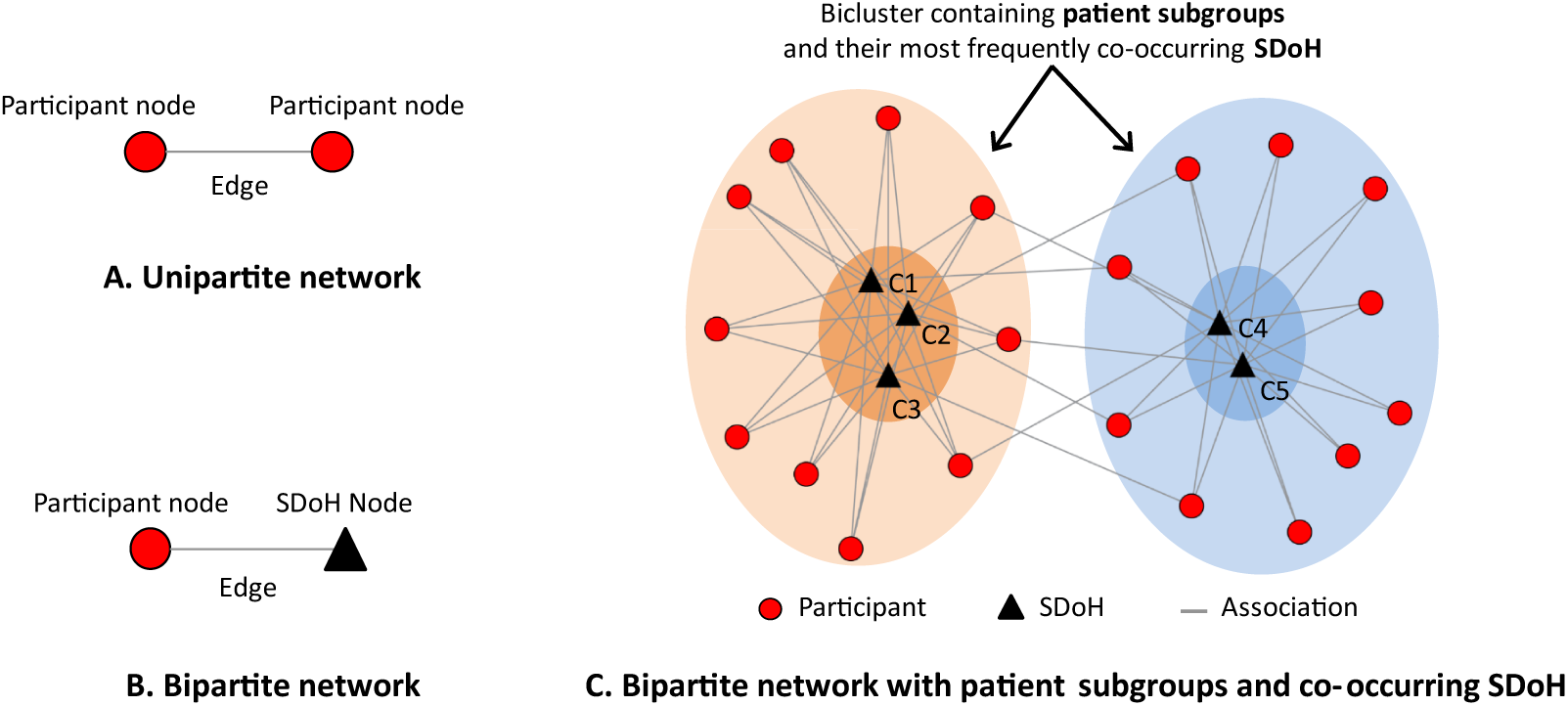
The distinction between a unipartite network (A), a bipartite network (B), and how the latter can be used to identify biclusters of participants and their most frequently co-occurring SDoH (C).

## Appendix B

SNOMED Codes Related to SDoH, and their Use in the Electronic Health Records of Participants in *All of Us*.

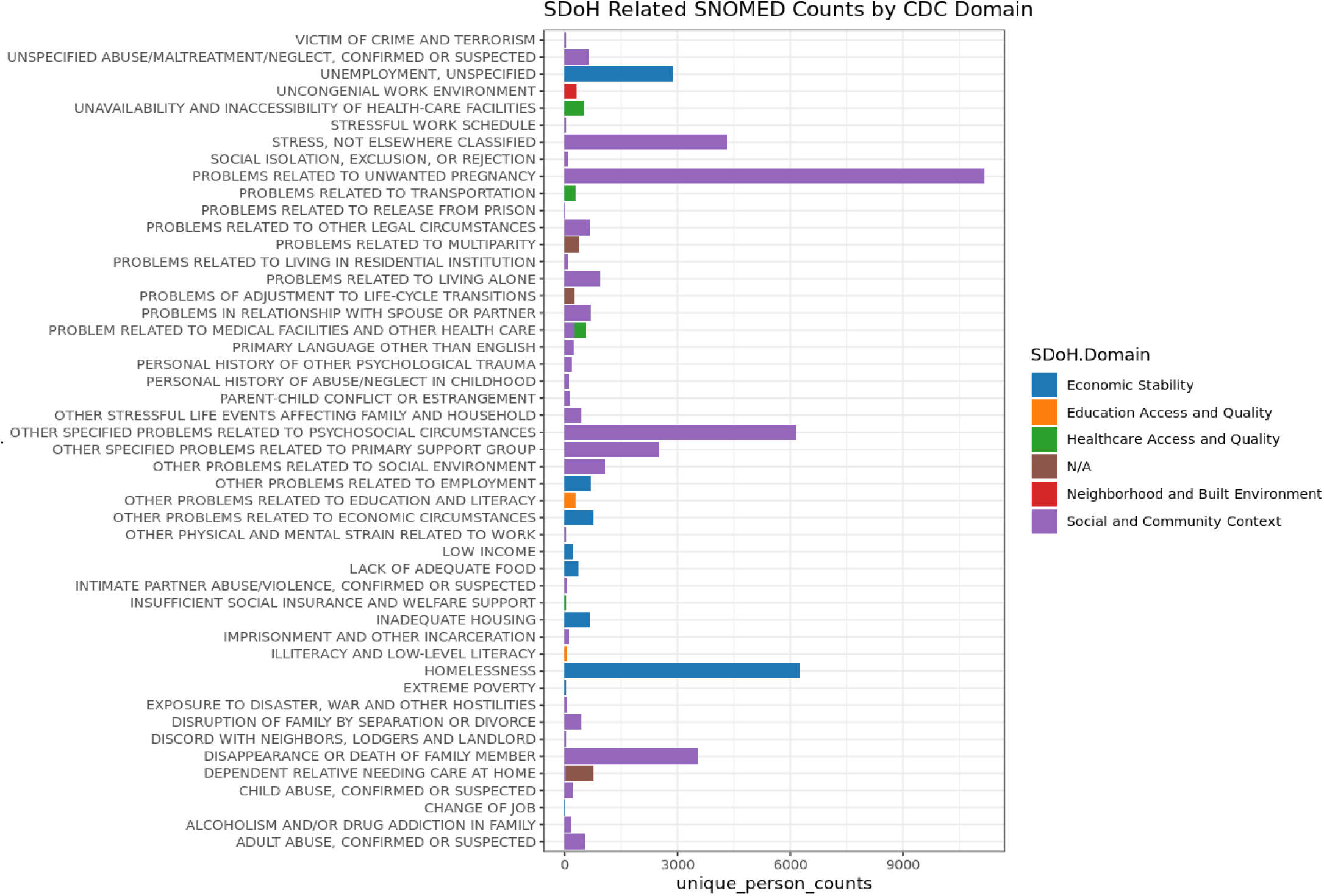

## Appendix C

Four *All of Us* surveys (Column-2), contained 110 SDoH questions (Column-3), that were abbreviated, negatively phrased (shown bolded) and reversed coded (shown in red) (Column-3), categorized into the five *HP-30* domains (Column-4 and shown by the five colors), and further categorized (boxes) by the expert panel into 18 factors (Column-5; *Delayed Medical Care* was used as an outcome).

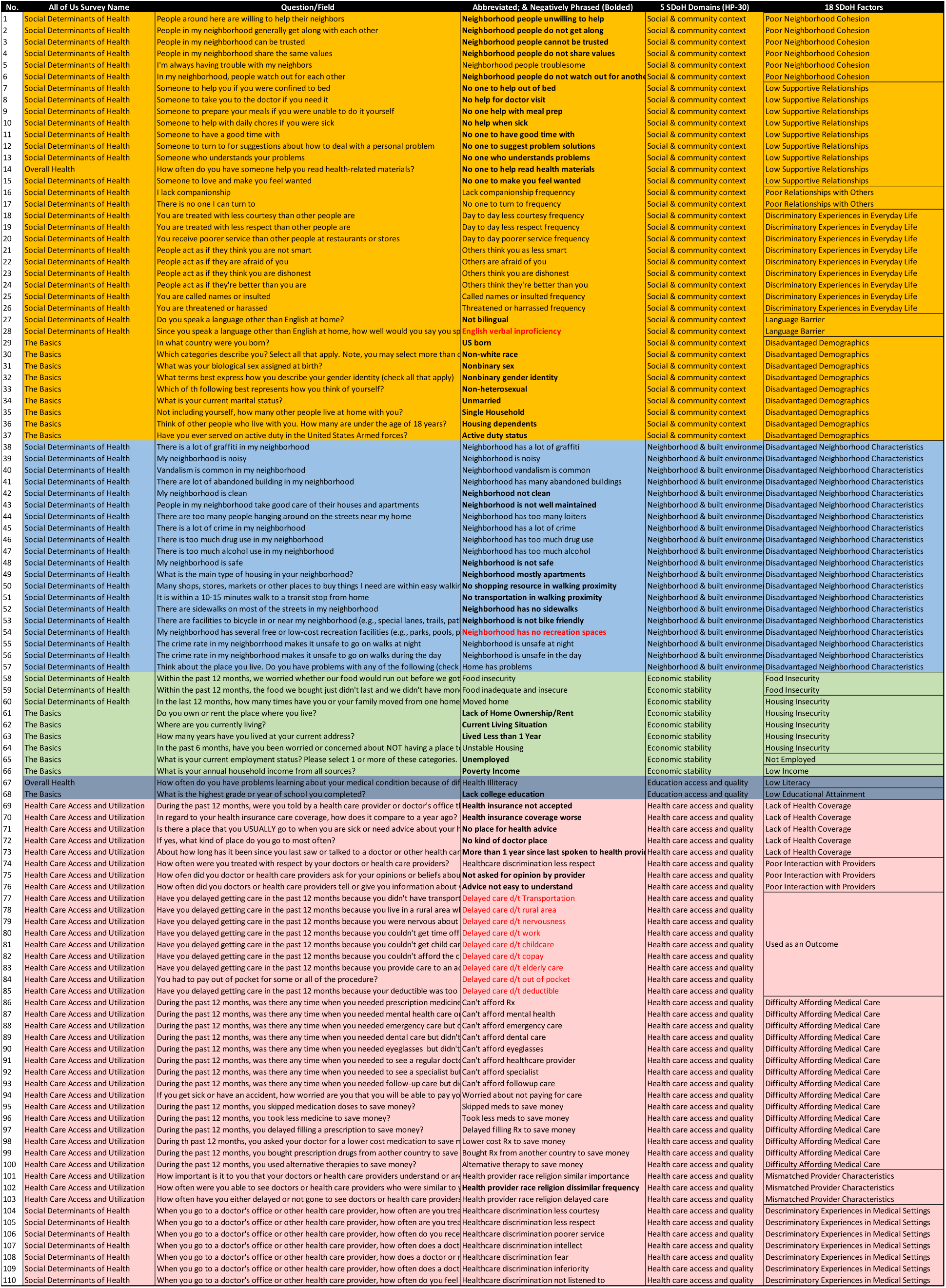

## Appendix D

Condition-specific cohort extraction for type II diabetes (T2DM), breast cancer, and coronary artery disease (CAD). **Figure 1**. Inclusion and exclusion criteria for selecting three condition-specific cohorts.

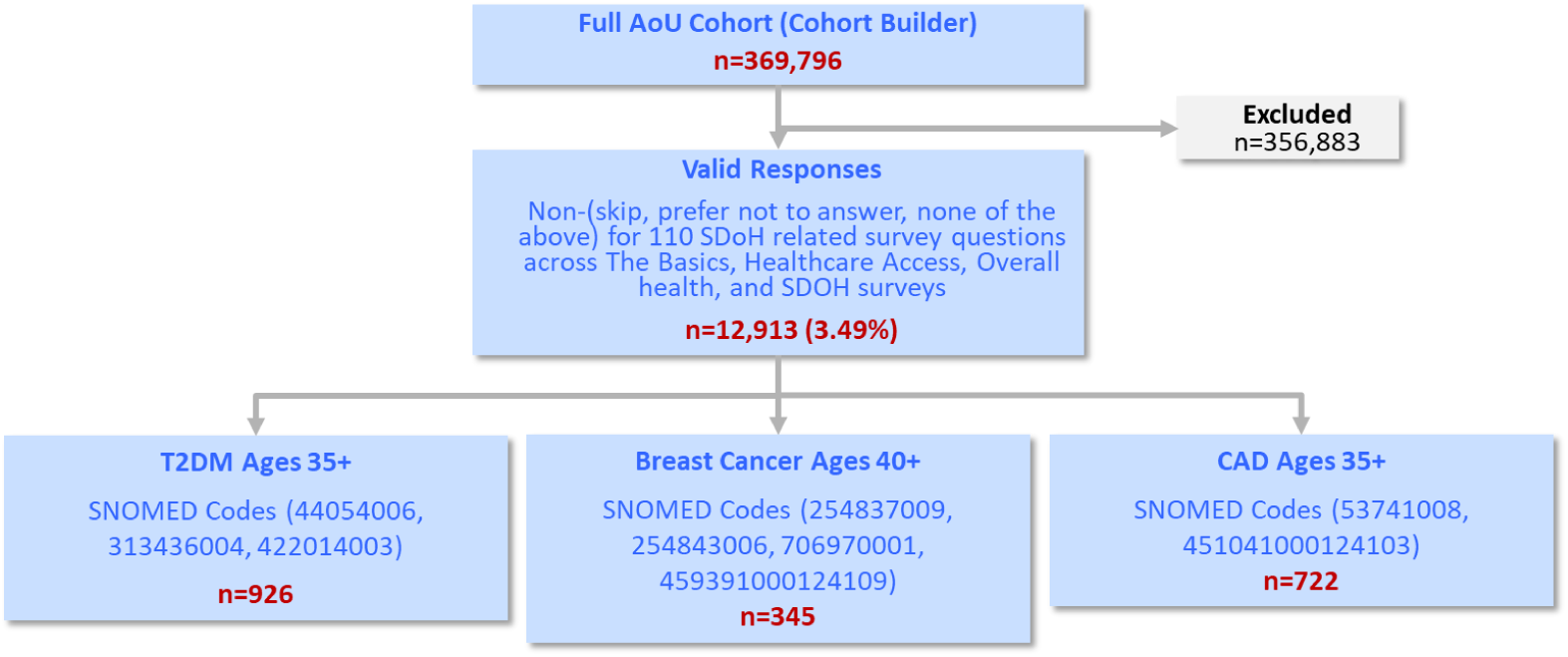

## Appendix E

Inverse Probability Weighting (IPW)We found significant differences in the demographic proportions between our cohort (n=12,913) consisting of participants with valid answers for all 110 SDoH questions, and the total *All of Us* data. To adjust for potential sample selection bias, we calculated inverse probability weights (IPW) using the *ipwpoint* function in the R package *ipw*.^66^ This function uses a logistic regression model to estimate the predicted probability of having valid responses on all SDoH variables based on age, sex, race, ethnicity, being born in the United States, currently employed, having a college degree or higher, health insurance, owning a home, and being married. We stabilized the weights according to the observed probability of being in our cohort. The resulting IPW weights were used as weights for the edges in the bipartite network.

